# Diminished antibody response against SARS-CoV-2 Omicron variant after third dose of mRNA vaccine in kidney transplant recipients

**DOI:** 10.1101/2022.01.03.22268649

**Authors:** Ayman Al Jurdi, Rodrigo B. Gassen, Thiago J. Borges, Isadora T. Lape, Leela Morena, Orhan Efe, Zhabiz Solhjou, Rania El Fekih, Christa Deban, Brigid Bohan, Vikram Pattanayak, Camille N. Kotton, Jamil R. Azzi, Leonardo V. Riella

## Abstract

**Background:** Available SARS-CoV-2 vaccines have reduced efficacy against the Omicron variant in immunocompetent individuals. Kidney transplant recipients (KTRs) have diminished antiviral responses to wild-type SARS-CoV-2 after vaccination, and data on antiviral responses to SARS-CoV-2 variants, including the Omicron variant, are limited.

**Methods:** We conducted a prospective, multi-center cohort study of 51 adult KTRs who received three doses of BNT162b2 or mRNA-1273. Blood and urine samples were collected before and four weeks after the third vaccine dose. The primary outcome was anti-viral antibody responses against wild-type and variants of SARS-CoV-2. Secondary objectives included occurrence of breakthrough SARS-CoV-2 infection and non-invasive monitoring for rejection using serum creatinine, proteinuria, donor-derived cell-free DNA and donor-specific antibodies. Sera from pre-pandemic healthy controls and KTRs were used for comparison.

**Results:** 67% of KTRs developed anti-wild-type spike antibodies after the third vaccine dose, similar to the Alpha (51%) and Beta (53%) variants, but higher than the Gamma (39%) and Delta (25%) variants. No KTRs had neutralizing responses to the Omicron variant before the third vaccine dose. After the third dose, fewer KTRs had neutralizing responses to the Omicron variant (12%) compared to wild-type (61%) and Delta (59%) variants. Three patients (6%) developed breakthrough SARS-CoV-2 infection at a median of 89 days. No KTRs developed allograft injury, *de novo* donor-specific antibodies or allograft rejection.

**Conclusion:** In KTRs, a third dose of mRNA vaccines increases antibody responses against wild-type and variants of SARS-CoV-2, while neutralizing responses to the Omicron variant remain markedly reduced.

## INTRODUCTION

Coronavirus disease (COVID-19) caused by severe acute respiratory coronavirus 2 (SARS-CoV-2) has resulted in over 800,000 deaths in the United States^1^ with the Delta (B.1.617.2) and more recently the Omicron (B.1.1.529) variants accounting for the majority of cases.^2^ Kidney transplant recipients (KTRs) are at increased risk of mortality following SARS-CoV-2 infection,^3^ and develop blunted antiviral responses following SARS-CoV-2 vaccination compared to non-transplant patients.^4–6^ Furthermore, the Delta and Omicron variants have been shown to be less sensitive to neutralizing antibodies from sera of vaccinated immunocompetent individuals,^7,8^ and similar data from vaccinated KTRs are limited.^9^ Therefore, in an era of clinically prevalent SARS-CoV-2 variants, evaluating antiviral responses to both wild-type (WT) and variants of SARS-CoV-2 in KTRs following SARS-CoV-2 mRNA vaccination is crucial. The aim of this study was to evaluate antiviral humoral responses against SARS-CoV-2 variants, including the Delta and Omicron variants following the third dose of SARS-CoV-2 mRNA vaccination in KTRs.

## MATERIALS AND METHODS

### Study design and patient recruitment

This is a prospective multicenter observational cohort study of the antiviral immune responses against SARS-CoV-2 variants following the third dose of SARS-CoV-2 mRNA vaccination in adult KTRs. Inclusion criteria were KTRs aged ≥18 years who were >3 months post-transplantation with stable allograft function and no rejection in the preceding six months. Full exclusion criteria are listed in supplementary materials. Subjects were enrolled consecutively.

Enrolled KTRs had received two doses of mRNA-1273 or BNT162b2 SARS-CoV-2 mRNA vaccines and were planned to receive a third dose of the same vaccine as per national guidelines. Participants had a baseline visit prior to the third vaccine dose and a follow-up visit one month following the third vaccine dose, where blood and urine samples were collected. Samples from fifteen pre-pandemic kidney transplant control patients and five pre-pandemic healthy controls were used for comparison.

### Study approval

The study was approved by the institutional review board at Mass General Brigham (IRB 2021P000043). All subjects signed written informed consent forms prior to enrollment in the study. The study was conducted in accordance with the Declaration of Helsinki and the Declaration of Istanbul. Data are reported in compliance with the STROBE statement reporting guidelines.

### Outcomes

The primary outcome was the development of antiviral humoral responses following the third vaccine dose as assessed by 1) measurement of antibodies directed against the spike protein and receptor-binding domain (RBD) of WT and Alpha (B.1.1.7), Beta (B.1.351), Gamma (P.1), Delta (B.1.617.2) and Omicron (B.1.1.529) variants of SARS-CoV-2 by a Luminex-based multiplex assay and by enzyme-linked immunosorbent assay (ELISA); and 2) Antibodies neutralization capacity against WT, Delta and Omicron variants of SARS-CoV-2 using a surrogate virus neutralization test.^10^ Secondary objectives included 1) the occurrence of any severe or grade 4 related adverse events; 2) development of breakthrough SARS-CoV-2 infection; and 3) monitoring for rejection using serum creatinine, urine protein-to-creatinine ratio, donor-derived cell-free DNA and donor-specific antibodies (DSAs). Details of sample processing and assays are described in supplementary materials and methods.

### Statistics

Continuous variables are presented as means (± standard deviation) or as medians (with interquartile ranges or full ranges) depending on normality of distribution. Categorical variables are presented as frequencies and percentages. Differences between paired samples were assessed using a paired t-test, a Wilcoxon matched-pairs signed rank test, a repeated measures ANOVA or Friedman test as appropriate. For comparisons between continuous variables between three or more groups, if testing reached statistical significance, then pairwise testing was performed to determine significant differences between groups, using Dunn’s correction to adjust for multiple comparisons. For categorical variables, the differences in proportions were calculated using a Pearson’s Chi squared test or Fisher’s exact test as appropriate. All tests used were two-sided and a two-sided α level of 0.05 was considered to be statistically significant. SPSS v24 (Chicago, IL) and GraphPad Prism v9.1.2 (San Diego, CA) were used for statistical analysis and creation of figures.

## RESULTS

### Patient characteristics

Fifty-one vaccinated KTRs were enrolled in the study. Baseline characteristics are shown in Table 1. Median age was 63 years, 43% were female and forty-eight (94%) patients received the BNT162b2 vaccine. The median time between the second and third vaccine doses was 187 days (IQR 181-193). Baseline samples were collected at a median of 5 days (IQR 0-18) prior to the third vaccine dose and post-vaccination samples were obtained at a median of 29 days (IQR 26-33) post-vaccination.

**Table 1.** Baseline characteristics of kidney transplant recipients.

| Baseline characteristic | n=51 |
| --- | --- |
| Age at enrollment (years), median (IQR) | 63 (54-69) |
| Months between transplantation and 1<sup>st</sup> vaccine dose, median (range) | 43 (4-443) |
| Female sex, n (%) | 23 (43) |
| Previous kidney transplant, n (%) |  |
| None | 47 (92) |
| One | 3 (6) |
| Two | 1 (2) |
| Cause of ESKD, n (%) |  |
| Glomerular disease | 21 (41) |
| Polycystic kidney disease | 9 (18) |
| Diabetic nephropathy | 5 (10) |
| Other genetic kidney disease | 5 (10) |
| Lithium toxicity | 2 (4) |
| Other or unknown | 9 (18) |
| Pre-transplant RRT, n (%) |  |
| None | 19 (37) |
| Hemodialysis | 27 (53) |
| Peritoneal dialysis | 5 (10) |
| Donor source, n (%) |  |
| Living related | 11 (22) |
| Living unrelated | 22 (43) |
| Deceased | 18 (35) |
| Cold ischemia time (hours), median (IQR) | 1.3 (0.8-12.1) |
| KDPI (%), median (IQR) | 57 (42-69) |
| HLA ABDR mismatches, median (IQR) | 4 (3-5) |
| Class I PRA (%), median (range) | 0 (0-69) |
| Class II PRA (%), median (range) | 0 (0-97) |
| Pre-transplant DSA, n (%) | 2 (4) |
| DSA at the time of vaccination, n (%) | 6 (12) |
| Induction immunosuppression, n (%) |  |
| Anti-thymocyte globulin | 28 (55) |
| Basiliximab | 16 (31) |
| Data not available | 7 (14) |
| Maintenance immunosuppression, n (%) |  |
| Calcineurin inhibitor |  |
| Cyclosporine | 1 (2) |
| Tacrolimus | 26 (51) |
| Trough level in ng/mL, median (IQR) | 5.7 (4.9-6.6) |
| mTOR inhibitor |  |
| Everolimus | 2 (4) |
| Sirolimus | 2 (4) |
| Belatacept | 22 (43) |
| Mycophenolate | 35 (69) |
| Total daily dose in mg, median (IQR) | 1,000 (875-1,000) |
| Azathioprine | 5 (10) |
| Total daily dose in mg, median (range) | 62.5 (50-100) |
| Prednisone | 42 (82) |
| Total daily dose in mg, median (IQR) | 5 (5-5) |
| CMV serostatus, n (%) |  |
| D+/R+ | 4 (8) |
| D+/R- | 7 (14) |
| D-/R+ | 5 (10) |
| D-/R- | 24 (46) |
| Not available | 11 (22) |
| EBV serostatus, n (%) |  |
| D+/R+ | 31 (61) |
| D+/R- | 4 (8) |
| D-/R+ | 3 (6) |
| D-/R- | 1 (2) |
| Not available | 12 (24) |
| Delayed graft function, n (%) | 10 (20) |
| History of allograft rejection, n (%) | 11 (22) |
| Months between most recent rejection and 1<sup>st</sup> vaccine dose, median (IQR) | 30 (9-51) |
| Serum creatinine (mg/dL), median (IQR) | 1.25 (1.05-1.59) |
| Estimated GFR (ml/min/1.73 m<sup>2</sup>), mean ± SD | 56 ± 17 |
| Urine protein to creatinine ratio (g/g), median (IQR) | 0.12 (0.07-0.36) |
| Donor-derived cell free DNA (%), median (IQR) | 0.15 (0.00-0.23) |
| Previous SARS-CoV-2 infection, n (%) | 4 (8) |
| mRNA vaccine received, n (%) |  |
| BNT162b2 | 48 (94) |
| mRNA-1273 | 3 (6) |
| Days between 1<sup>st</sup> and 2<sup>nd</sup> vaccine dose, median (IQR) | 21 (21-21) |
| Days between 2<sup>nd</sup> and 3<sup>rd</sup> vaccine dose, median (IQR) | 186 (181-193) |
| ALC at time of 3<sup>rd</sup> vaccine dose, cells/mm<sup>3</sup>, median (IQR) | 1,395 (930-1,983) |
| ACE inhibitor or ARB use, n (%) | 11 (22) |

ACE: Angiotensin converting enzyme. ALC: Absolute lymphocyte count. ARB: Angiotensin receptor blocker. CMV: Cytomegalovirus. EBV: Epstein-Barr virus. ESKD: end-stage kidney disease. GFR: glomerular filtration rate. HLA: human leukocyte antigen. KDPI: kidney donor profile index. mTOR: mammalian target of rapamycin. PRA: panel reactive antibodies. RRT: renal replacement therapy. SD: standard deviation.

### Anti-spike antibody measurement

To evaluate antibody-mediated antiviral responses, we measured antibodies directed against the nucleocapsid (NC), spike trimer, S1 and RBD regions of the WT SARS-CoV-2 prior to and following the third vaccine dose in KTRs using a Luminex-based multiplex assay. After the third vaccine dose, there was no significant change in the MFI ratios of anti-NC antibodies (p=0.278) but there was a significant increase in the MFI ratios for antibodies directed against the spike trimer, S1 and RBD regions (p<0.001 for all, Fig. 1A). Using the recommended assay positivity threshold, all KTRs had a negative anti-NC antibody result before and after the third vaccine dose. The percentage of KTRs with anti-WT spike antibodies increased from 29% to 67% (p<0.001), anti-WT S1 antibodies increased from 10% to 47% (p<0.001) and anti-WT RBD antibodies increased from 12% to 45% (p<0.001) after the third vaccine dose. In comparison, all pre-pandemic healthy controls (HCs) and pre-pandemic kidney transplant patient controls (KCs) had negative results for antibodies against all four WT SARS-CoV-2 antigens (Fig. 1B). After the third vaccine dose, there was a significant increase in the MFI ratios of anti-spike antibodies directed against the Alpha, Beta, Gamma and Delta variants (p<0.001 for all, Fig. 1C), while all pre-pandemic HCs and KCs had undetectable antibodies against those variants (Fig. 1D). When evaluating differences in anti-spike antibody responses to SARS-CoV-2 variants, we found that prior to the third vaccine dose a higher percentage of KTRs (29%) had anti-spike antibodies against the WT virus compared to the Alpha (12%, p=0.028), Gamma (6%, p=0.002) and Delta (2%, p<0.001) variants but not the Beta variant (22%, p=0.364, Fig. 1E). After the third vaccine dose, a higher percentage of KTRs (67%) had anti-spike antibodies against the WT virus compared to the Gamma (39%, p=0.006) and Delta (25%, p<0.001) variants but not the Alpha (51%, p=0.108) or Beta variants (53%, p=0.158, Fig. 1F).

**Figure 1.**
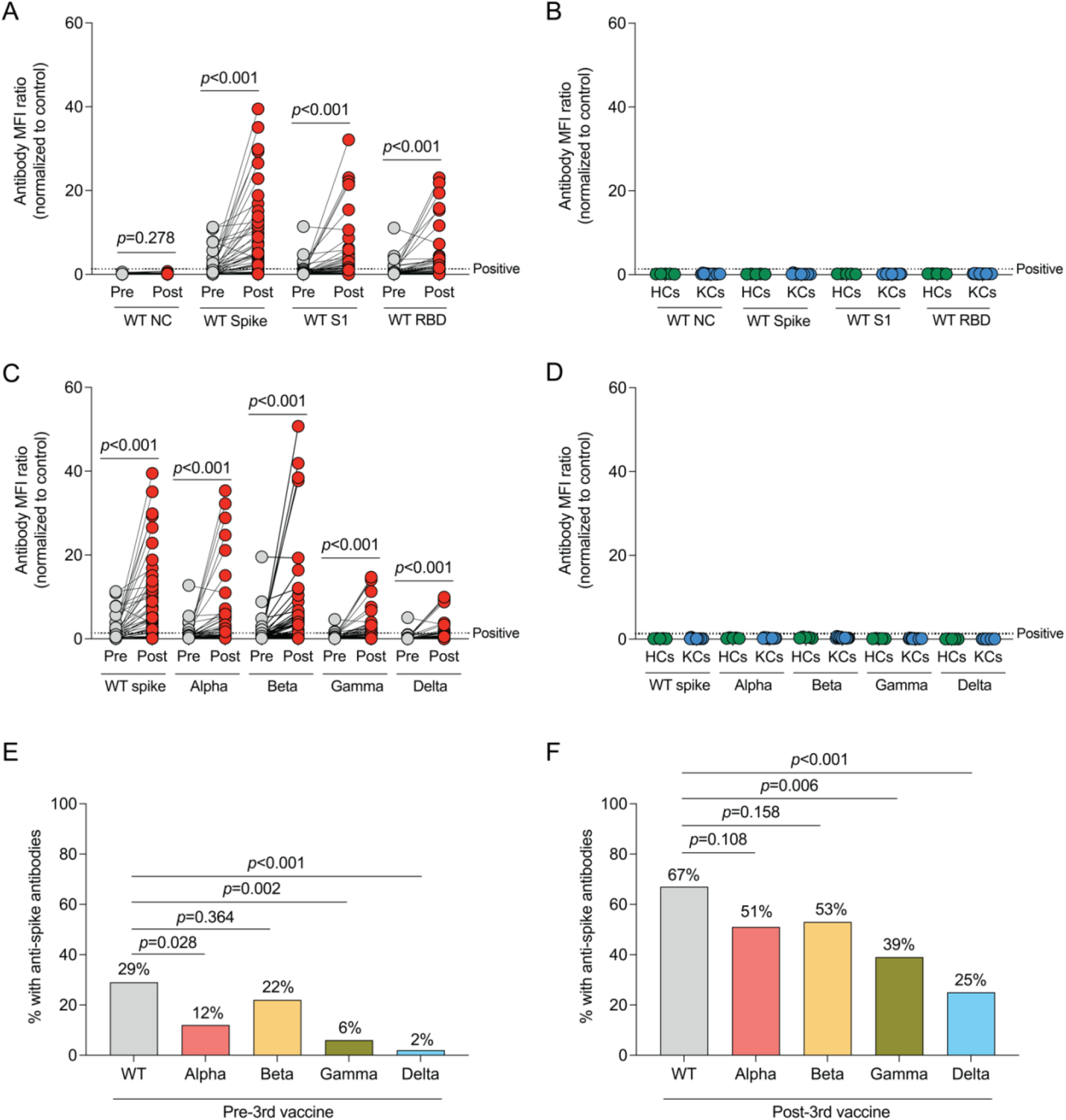
Increase in anti-spike antibody levels after the third dose of SARS-CoV-2 mRNA vaccine in kidney transplant recipients (KTRs). **(A)** Antibody MFI ratios to wild-type (WT) SARS-CoV-2 antigens before and after the third vaccine dose in KTRs (n=51) and **(B)** in pre-pre-pandemic healthy controls (HCs, n=5) and pre-pandemic kidney transplant control patients (KCs, n=15) measured by Luminex-based multiplex assay. **(C)** Antibody MFI ratios for anti-spike antibodies of WT and variants of SARS-CoV-2 before and after the third vaccine dose in KTRs and **(D)** pre-pandemic HCs and KCs. Horizontal lines indicate positivity threshold for assay. **(E)** Proportion of KTRs with anti-spike antibodies against WT and variants of SARS-CoV-2 before and **(F)** after the third vaccine dose. (**A, C**) Statistic by Wilcoxon matched-pairs signed rank test. **(E, F)** Statistic by Chi-square test. NC: Nucleocapsid. RBD: Receptor-binding domain.

### Anti-RBD antibody measurement against Omicron variant and its neutralization capacity

Since the multiplex assay lacked assessment of antibodies directed against the new Omicron variant, we evaluated anti-RBD antibodies against the WT, Delta and Omicron variants using ELISA. After the third vaccine dose, there was a significant increase in anti-RBD antibody levels for the WT (p<0.001), Delta (p<0.001) and Omicron (p<0.001, Fig. 2A) variants. In comparison, pre-pandemic HCs and KCs had no detectable antibodies against the RBD proteins of the WT, Delta and Omicron variants (Fig. 2B). After the third vaccine dose in KTRs, anti-RBD antibody levels were higher for the WT compared to the Omicron variant (p<0.001) and similar between WT and the Delta variant (p=0.142). There was a trend towards lower anti-RBD antibody levels for the Omicron compared to the Delta variant, but this did not reach statistical significance (p=0.110).

**Figure 2.**
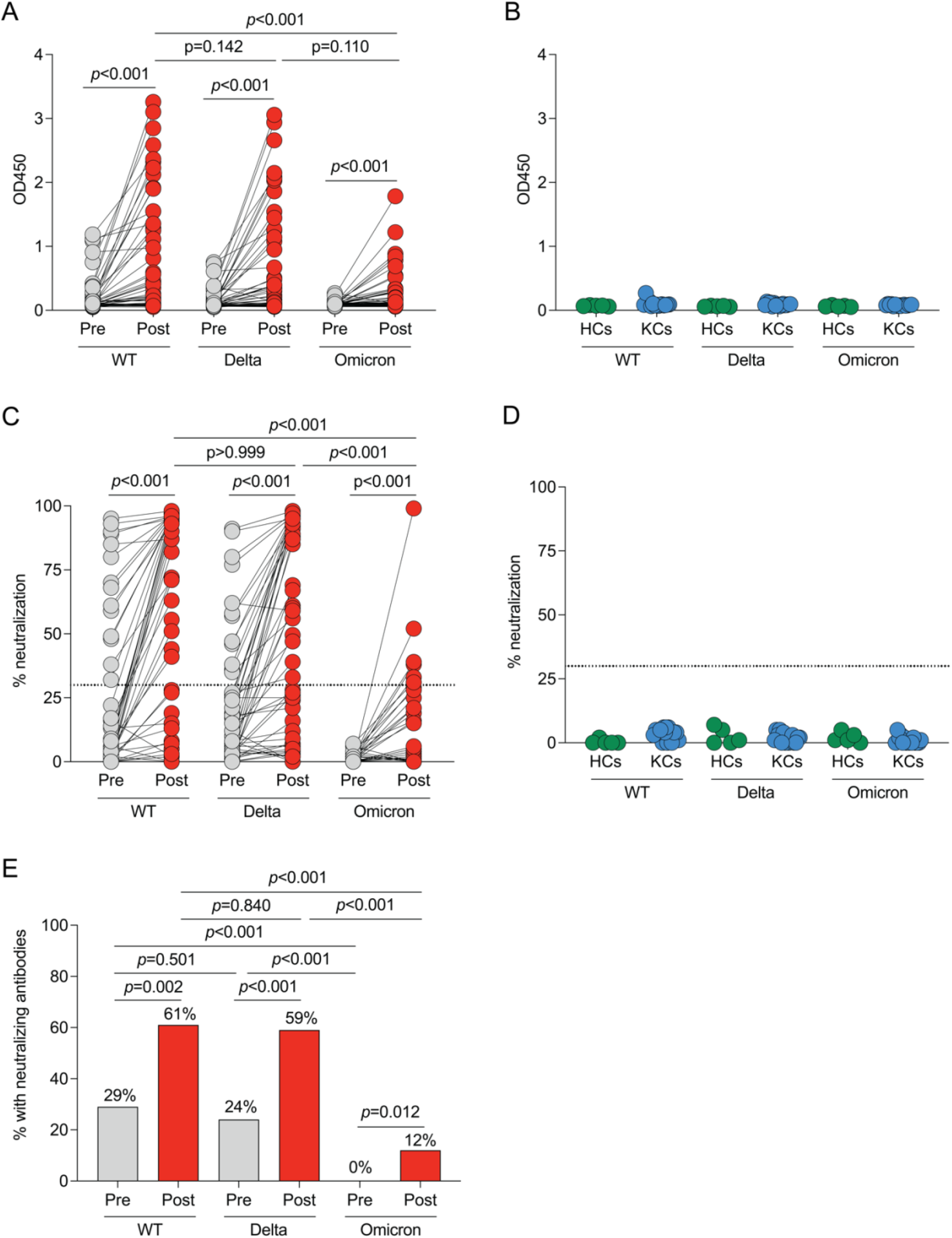
Poor anti-receptor binding domain (RBD) antibody and neutralization responses to the Omicron variant before and after the third dose of SARS-CoV-2 mRNA vaccine in kidney transplant recipients (KTRs). **(A)** Anti-RBD antibody OD450 values to wild-type (WT), Delta (B.1.617.2) and Omicron (B.1.1.529) variants of SARS-CoV-2 before and after the third vaccine dose in KTRs (n=51), and **(B)** in pre-pandemic healthy controls (HCs, n=5) and pre-pandemic kidney transplant control patients (KCs, n=15) measured by enzyme-linked immunosorbent assay. **(C)** Percentage of neutralization to WT, Delta and Omicron variants of SARS-CoV-2 before and after the third vaccine dose in KTRs (n=51) and **(D)** in pre-pandemic HCs (n=5) and KCs (n=15) measured by surrogate virus neutralization test. Horizontal lines indicate positivity threshold for assay. **(E)** Proportion of KTRs with a positive neutralization response before and after the third vaccine dose in KTRs. (**A, C**) Statistic by Friedman test with Dunn’s correction for multiple comparisons. **(E)** Statistic by Chi-square test.

We then evaluated neutralizing antibody responses using a surrogate virus neutralization test (SVNT).^10^ We found a significant increase in the percentage of neutralization against the WT (p<0.001), Delta (p<0.001) and Omicron variants (p<0.001, Fig. 2C) after the third vaccine dose. However, the percentage of neutralization was significantly lower for the Omicron variant compared to the WT and Delta variants (p<0.001). In comparison, there was no significant neutralizing response against the WT, Delta and Omicron variants in pre-pandemic HCs and KCs (Fig. 2D).

In addition, we found that the percentage of KTRs who had a neutralizing response increased from 29% to 61% for WT virus (p=0.002), from 24% to 59% for the Delta variant (p<0.001), and from 0% to 12% for the Omicron variant following the third vaccine dose (p=0.012, Fig. 2E). Prior to the third vaccine dose, fewer KTRs had a neutralizing response against the Omicron variant, compared to the WT (p<0.001) and Delta (p<0.001) variants, but there was no difference between the proportion of KTRs with neutralizing responses to the WT vs Delta variants (p=0.501). Similarly, after the third vaccine dose, fewer KTRs had a neutralizing response against the Omicron variant, compared to the WT (p<0.001) and Delta (p<0.001) variants, but there was no difference between the proportion of KTRs with neutralizing responses to the WT vs Delta variants (p=0.840). Moreover, the KTRs with neutralizing responses to the Delta variant but without anti-Delta spike antibodies by the multiplex assay had anti-WT spike antibodies.

### Breakthrough infection after vaccination

At a median follow-up of 89 days (IQR 78-100) after the third vaccine dose, three patients (6%) developed symptomatic SARS-CoV-2 infection (Table 2). None had neutralizing responses to the Omicron variant and two did not have neutralizing responses to the WT and Delta variant. All had mild symptoms and were treated with monoclonal antibodies in the outpatient setting. All recovered without requiring hospital admission and none received dexamethasone or remdesivir.

**Table 2.**
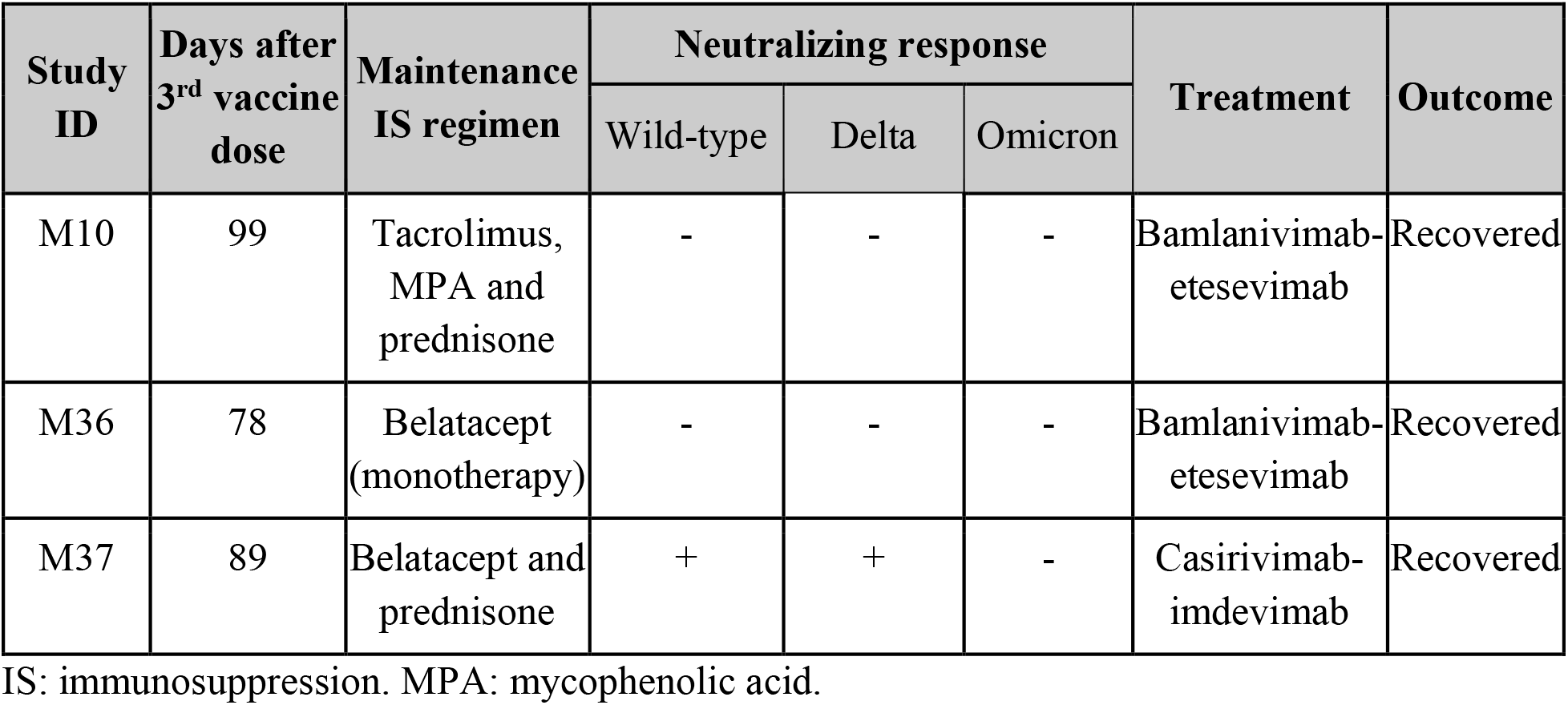
Breakthrough SARS-CoV-2 infection after third dose of vaccination.

| Study ID | Days after 3<sup>rd</sup> vaccine dose | Maintenance IS regimen | Neutralizing response |  |  | Treatment | Outcome |
| --- | --- | --- | --- | --- | --- | --- | --- |
|  |  |  | Wild-type | Delta | Omicron |  |  |
| M10 | 99 | Tacrolimus, MPA and prednisone | - | - | - | Bamlanivimab-etesevimab | Recovered |
| M36 | 78 | Belatacept (monotherapy) | - | - | - | Bamlanivimab-etesevimab | Recovered |
| M37 | 89 | Belatacept and prednisone | + | + | - | Casirivimab-imdevimab | Recovered |

IS: immunosuppression. MPA: mycophenolic acid.

### Non-invasive rejection monitoring, allograft rejection and adverse events

No patients experienced severe adverse events post-vaccination. At a median of 29 days (IQR 26-32) after the third vaccine dose, no KTRs developed *de novo* DSAs and there was no significant change in serum creatinine (p=0.212), urine protein-to-creatinine ratio (p=0.257), or donor-derived cell-free DNA levels (p=0.434, Table S1) compared to pre-vaccination. At a median follow-up of 89 days (IQR 78-100) after the third vaccine dose, no patients developed allograft rejection.

## DISCUSSION

This study is one of the first to characterize antiviral humoral responses against SARS-CoV-2 variants, including the Omicron variant, after a third dose of SARS-CoV-2 mRNA vaccines in KTRs. Using a Luminex-based multiplex assay, we found that two thirds of KTRs had anti-WT spike antibodies after the third vaccine dose, similar to previous reports.^11–16^ Despite an increase in the proportion of KTRs with anti-spike antibodies against WT SARS-CoV-2 and variants after the third vaccine dose, fewer KTRs had anti-spike antibodies against the Gamma and Delta variants compared to the WT, Alpha and Beta variants. Using an ELISA we developed in our laboratory, we found an increase in anti-RBD antibody levels against the WT, Delta and Omicron variants after the third vaccine dose, although anti-RBD antibody levels were significantly lower for the Omicron variant compared to WT virus. To assess neutralization capacity of antibody responses, we used a SVNT, which has been shown to be highly correlated with live virus neutralization assays.^10^ We found an increase in the percentage of neutralization against the WT, Delta and Omicron variants after the third vaccine dose as has been reported in immunocompetent individuals.^17,18^ About half of KTRs had neutralizing responses to the WT and Delta variants after third vaccine dose, consistent with what has been reported previously in KTRs.^19^ The presence of Delta variant neutralization in KTRs with anti-WT but not anti-Delta spike antibodies suggests cross-reactivity of anti-WT spike antibodies with the Delta variant. Concerningly, no KTRs had neutralizing responses to the Omicron variant prior to the third vaccine dose and only 12% had neutralizing responses to the Omicron variant after the third vaccine dose. The Omicron variant’s ability to escape neutralization compared to WT SARS-CoV-2 in our study is consistent with data from vaccinated immunocompetent individuals^8,17,20,21^ and is likely due to its highly mutated RBD region, which has 15 mutations compared to WT RBD.^22–24^ It is worth noting that a limitation of the SVNT is that it is not able to measure neutralizing antibodies directed against non-RBD regions of the spike protein as it only measures RBD-ACE2 interactions.

We were thus able to provide a detailed characterization of antibody responses to SARS-CoV-2 variants in KTRs, including the Omicron variant, and to evaluate the alloimmune safety of a third vaccine dose and found no evidence of allograft injury, *de novo* DSA development and no episodes of allograft rejection. This is consistent with what has been reported after two^25–27^ and three vaccine doses^11,12,14^ in solid organ transplant recipients. Our study has limitations, including its small sample size, observational design, lack of a control group of KTRs who did not receive a third vaccine dose, and lack of assessment of the cellular response to vaccination. Further studies evaluating the cellular response to the Omicron variant and the implications of the reduced neutralization ability with regards to risk and severity of infections in KTRs are needed.

In summary, we found that a third dose of SARS-CoV-2 mRNA vaccination in KTRs was associated with an increased antiviral antibody response against WT and variants of SARS-CoV-2, and while the neutralizing responses to the Omicron variant increased in some, overall they remained markedly diminished. Strategies designed to improve antiviral immune responses to the Omicron and future variants, such as with additional homologous^28^ or heterologous^29^ vaccine doses or use of higher vaccine doses, are also needed in this vulnerable high-risk group, in particular with the surge of Omicron as the predominant variant worldwide.

## Data Availability

Data to support the findings in the study are available from the corresponding author upon reasonable request.

## Author contributions

Conceptualization: AAJ, RBG, TJB, ZS, JRA, LVR

Data curation: AAJ, LM, OE, ZS, RE

Formal analysis: AAJ, RBG, TJB, ITL, BB, VP, LVR

Funding acquisition: AAJ, JRA, LVR

Investigation: AAJ, RBG, TJB, ZS, RE, CD, ITL, BB, CK, LVR

Methodology: AAJ, RBG, TJB, ITL, OE, BB, VP, CK, JRA, LVR

Project administration: AAJ, ZS, RE, CD, LM, ITL, OE, VP, JRA, LVR

Validation: AAJ, ITL

Visualization: AAJ, RBG, TJB

Supervision: AAJ, ITL, BB, LVR

Writing-original draft: AAJ

Writing-review & editing: AAJ, RBG, TJB, ITL, CK, JRA, LVR

## Acknowledgments

We would like to thank the staff of the transplant clinic for their assistance in conducting this study.

