## Supplementary materials and tables for "Diminished antibody response against SARS-CoV-2 Omicron variant after third dose of mRNA vaccine in kidney transplant recipients"

**Supplementary Material**

Table of contents:

- Supplementary tables: page 2
- Supplementary materials and methods: pages 3-76
- References: pages 7

### Supplementary tables

Table S1. Allograft status before and four weeks after third SARS-CoV-2 mRNA vaccine dose.

| Biomarker | Before third dose | After dose | p value |
|---|---|---|---|
| Serum creatinine (mg/dL), median (IQR) | 1.25 (1.05-1.59) | 1.28 (1.05-1.49) | 0.212† |
| UPCR (g/g), median (IQR) | 0.12 (0.07-0.36) | 0.13 (0.08-0.37) | 0.257† |
| ddcfDNA (%), median (IQR) | 0.15 (0.00-0.23) | 0.15 (0.00-0.26) | 0.434† |

ddcfDNA: donor-derived cell-free DNA. IQR: interquartile range. UPCR: urine protein-to-creatinine ratio. †Statistic by Wilcoxon matched-pairs signed rank test.

**Supplementary materials and methods:**

Exclusion criteria:

- Age<18 years of age
- Unstable allograft function (>20% variation in last two eGFR values measured at least one week apart)
- Cellular or antibody-mediated rejection during the previous six months
- Multi-organ transplantation
- Pregnant or lactating females
- Allergy to any component of mRNA-1273 or mRNA-BNT162b2 vaccines
- Investigational drug use within 30 days of enrollment
- Receipt of non-live vaccine with 2 weeks or live viral vaccine within 4 weeks of SARS-CoV-2 vaccination
- Acute or chronic illness at the time of vaccination which in the opinion of the investigator will alter immune response (e.g. human immunodeficiency virus infection, primary immunodeficiency disease, disseminated or untreated malignancy, or systemic infection).

Sample collection and processing

Blood and urine samples were collected from KTRs prior to and one month after the third vaccine dose. Blood and urine samples were sent to the clinical lab (for serum creatinine and urine protein-to-creatinine ratio quantification), our research laboratories (for anti-HLA antibody, anti-viral antibody immune assays) and CareDx, Inc. (for donor-derived cell-free DNA levels).

Serum and plasma were obtained from peripheral blood by centrifuging for 15 minutes at 2,500 RPM at room temperature then stored in cryogenic tubes at -80°C.

Antibody quantification by Luminex-based multiplex assay:

Total antibodies (IgM, IgA and IgG) directed against multiple WT SARS-CoV-2 antigens (spike protein trimer, S1 region, receptor-binding domain [RBD] region and nucleocapsid [NC]) and the spike protein from multiple SARS-CoV-2 variants (alpha [B.1.1.7], Beta [B.1.351], Gamma [P.1] and Delta [B.1.617.2]) were measured pre- and post-vaccination using the Coronavirus Ig Total Human 15-Plex ProcartaPlex™ Panel (Invitrogen™, catalog no. PPX-15-MXFVM2D). Capture beads were added to each well and then controls and samples (in a 1:1,000 dilution) were added. The plate was incubated for 2 hours at room temperature, then washed and detection antibody was added to each well. After a 30-minute incubation at room temperature, the plate was read using MAGPIX system (Luminex, Austin, TX) and the data was analyzed using xPONENT software (Luminex, Austin, TX). As indicated by the manufacturer, a positive result was defined as a sample to low-control MFI ratio above 1.3. An indeterminate result was defined as an MFI ratio between 1.0 and 1.3, while a MFI ratio <1.0 was defined as a negative result.

Antibody quantification by ELISA:

IgG antibodies directed against the RBD of WT, Delta (B.1.617.2) and Omicron (B.1.1.529) variants of SARS-CoV-2 were measured using an enzyme-linked immunosorbent assay (ELISA). Briefly, 96-well flat-bottom plates were coated with 100 µL of RBD protein for the respective variants (SinoBiological, catalog no. 40592-V08H, 40592-V08H90, and 40592-V08H121) at a concentration of 1 µg/mL. The plates were incubated overnight, washed with washing buffer and blocking solution (Thermo Scientific, catalog no. 37520) was added for 1 hour at room temperature (RT). The plates were washed then 100 µL of serum (diluted 1:3,000 in sample diluent solution, R&D Systems, catalog no. DY007) were added to each well and the plates were incubated for 2 hours at RT. The plates were then washed, 100 µL of anti-human IgG secondary antibody (BioLegend, catalog no. 410902, diluted 1:10,000) was added, and the plate was incubated for 1 hour at RT. 3,3',5,5'-Tetramethylbenzidine (TMB, ThermoFisher, catalog no. N301) was added and the plate was incubated for 15 minutes at RT. Finally, a stop solution (R&D Systems, catalog no. DY007) was added, and the plate was read at 450nm using SpectraMax iD3 microplate reader (Molecular Devices, San Jose, CA). Results are reported as OD450.

Antibody neutralizing capacity:

We evaluated the neutralizing function of antibodies using a surrogate virus neutralization test (GenScript cPass kit, catalog no. L00847-A), which has been shown to correlate well with conventional live virus neutralization test.[1] Patient sera (diluted 1:10) were incubated at 37°C for 30 minutes with horseradish peroxidase-conjugated recombinant SARS-CoV-2 WT, Delta variant RBD fragments (HRP-RBD, GenScript catalog no. Z03614-20) or Omicron variant RBD fragments (SinoBiological catalog no. 40592-V49H7-B combined with ThermoFisher catalog no. 434423) in a 1:1 ratio. The Delta variant peptide had L452R and T478K mutations. The Omicron variant peptide had G339D, S371L, S373P, S375F, K417N, N440K, G446S, S477N, T478K, E484A, Q493R, G496S, Q498R, N501Y, Y505H, T547K mutations. The mix of sera and HRP-RBD was added to each well of a capture plate pre-coated with human angiotensin converting enzyme 2 protein (ACE2) then incubated at 37°C for 15 minutes. Neutralizing antibodies form complexes with HRP-RBD that remain in the supernatant are removed with washing, while nonneutralizing antibodies-HRP-RBD complexes and unbound HRP-RBD bind to ACE2 and are captured on the plate. After washing, TMB solution was added, and the plate was incubated in the dark for 15 minutes at room temperature. Finally, a stop solution was added, and the plate was read at 450nm using SpectraMax iD3 microplate reader (Molecular Devices, San Jose, CA). The absorbance of the sample is inversely related to the concentration of neutralizing antibody. As indicated by the manufacturer, inhibition of ≥30% was considered a positive result for neutralization.

<u>Donor-derived cell-free DNA assay:</u>

Circulating ddcfDNA levels were measured as per published AlloSure® protocol (CareDx, Inc., Brisbane, CA).[2,3] Briefly, duplicate samples of venous blood were collected using Streck cell-free DNA BCT® tubes then shipped to CareDx, Inc. laboratories (Brisbane, CA), where plasma was separated by centrifugation then cell-free DNA was extracted using the circulating nucleic acid kit (Qiagen, cat no. 55114) following manufacturer's instructions. Plasma ddcfDNA levels were measured using a next-generation sequencing assay utilizing 266 single nucleotide polymorphisms, which allows quantification of ddcfDNA without requiring genotyping of the donor or recipient. Results are reported as percentage of total circulating cell-free DNA. Percentages above 0.5-1.0% are associated with an increased risk of allograft rejection.[3,4] Fifty one out of fifty eight patients had Streck cell-free DNA BCT® tubes collected for ddcfDNA measurement.

<u>Anti-human leukocyte antigen antibody assays</u>

Screening for anti-human leukocyte antigen (HLA) antibodies was performed at 3 months post-vaccination using mixed class I & II kit (One Lambda, catalog no. LSM12). Briefly, patient sera and negative control sera (One Lambda, catalog no. LS-NC) were added to HLA-coated beads and incubated for 30 minutes at room temperature. After washing, PE-conjugated goat anti-human IgG secondary antibody (One Lambda, catalog no. LS-AB2) was added then incubated for 30 minutes. The results were read using LABScan3D™ (One Lambda, Los Angeles, CA) and analyzed using HLA Fusion™ software (One Lambda, Los Angeles, CA). A sample to negative control serum mean fluorescent intensity (MFI) ratio >3.5 was considered positive per our histocompatibility lab's standards. Patients with a positive anti-HLA antibody screen at month 3 then had anti-HLA antibody class I and II testing using single-antigen beads (LABScreen™ Single Antigen Class I – Combi, catalog no. LS1A04 and Class II – Group 1, catalog no. LS2A01, One Lambda) with an identical protocol to determine which anti-HLA antibodies were present to determine if they were donor-specific. Baseline (pre-vaccination) sera were then tested to determine if DSAs had been present prior to vaccination or were *de novo* (defined as new DSAs with MFI >1,000).
